# Sex-modulated association between thyroid stimulating hormone and informant-perceived anxiety in non-depressed older adults: prediction models and relevant cutoff value

**DOI:** 10.1101/2024.07.26.24311073

**Authors:** Asma Hallab, Alzheimer’s Disease Neuroimaging Initiative

## Abstract

**Highlights:**

- Lower TSH levels predicted higher odds of anxiety in non-depressed older adults.
- The association between TSH and anxiety was significant in older males but not in older females.
- TSH level corresponding to 2.4 µIU/mL was a significant cutoff value in this association, under which thyroid function predicted significantly higher odds of anxiety in older males.
- Only in older males, but not older females, TSH levels were significantly lower in those with anxiety than in those without.

**Introduction:** The aim of this study was to assess the association between thyroid function and perceived anxiety in non-depressed older adults.

**Methods:** Non-depressed Alzheimer’s Disease Neuroimaging Initiative participants with complete Thyroid Stimulating Hormone (TSH) and neuropsychiatric inventory (NPI/NPI-Q) were included. The association between anxiety and thyroid function was assessed by logistic regression and sex stratification. Restricted cubic splines were applied to evaluate non-linearity in the association.

**Results:** The median age of 2,114 eligible participants was 73 years (68-78), 1,117 (52.84%) were males, and the median TSH was 1.69µIU/mL. There was a significant association between TSH and informant-perceived anxiety in the total study population (OR_Model1_=0.86, 95%CI 0.76-0.97, p=0.011), even after adjusting for bio-demographical (adj.OR_Model2_=0.85, 95%CI 0.75-0.96, p=0.007), and socio-cognitive confounders (adj.OR_Model3_=0.84, 95%CI 0.73-0.96, p=0.009). Sex-stratification showed similar significant results in all models only in males (OR_Model1-male_=0.71, 95%CI: 0.58-0.85, _pModel1-male_<0.001). In the general population and males, a TSH value of 2.4µIU/dL was a significant cutoff under which anxiety odds were significantly high, even after adjusting for confounders.

**Conclusions:** The sex-dependent association between TSH levels and perceived anxiety in non-depressed older adults is a novel finding that has to be further explored for a better understanding of the underlying neurobehavioral biology.

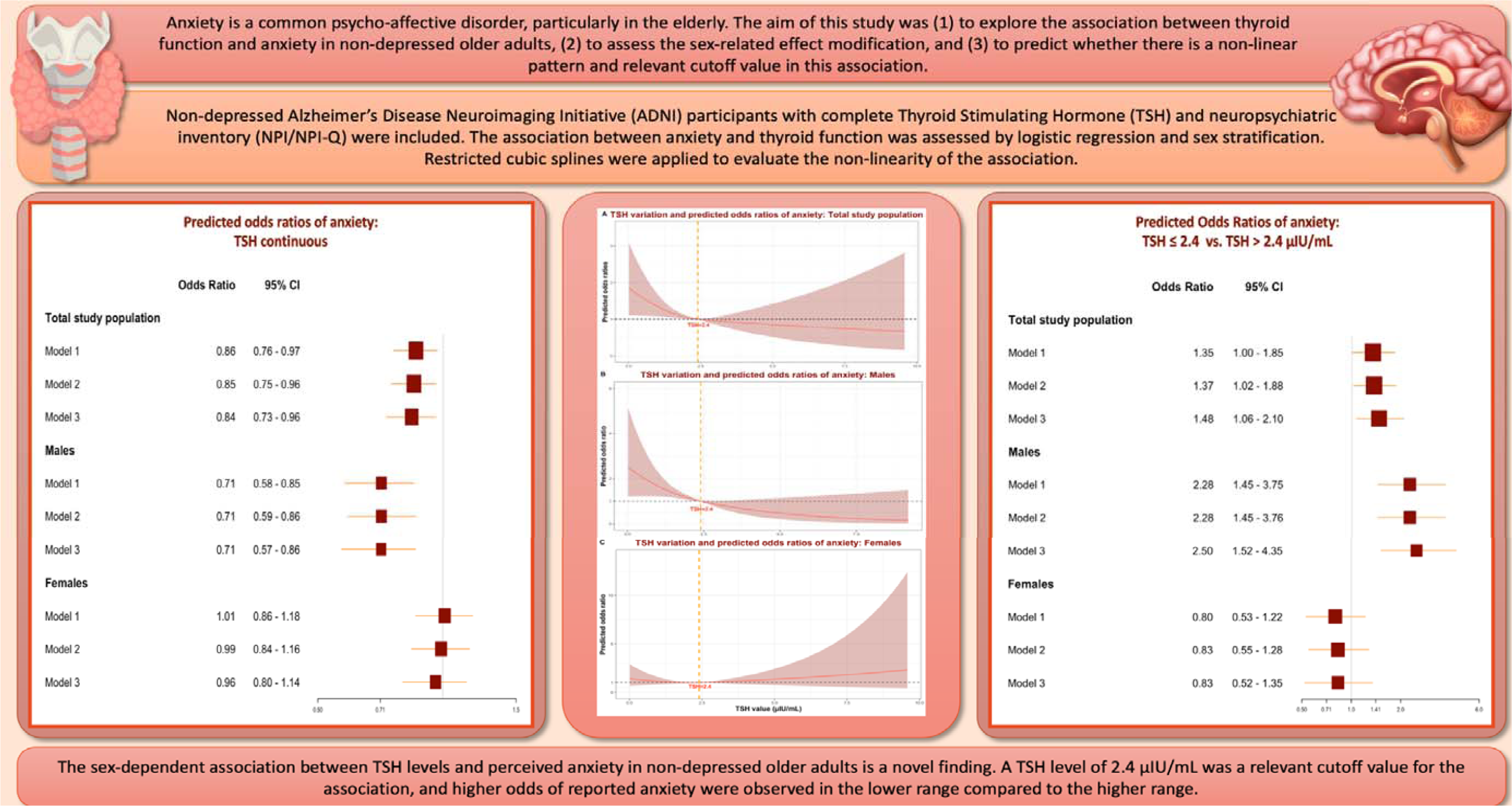

## 1. Introduction

Anxiety is a common psycho-affective disorder including generalized anxiety disorders, social anxiety disorders, panic disorders, and specific phobias. (1) It is largely diagnosed across different age groups, (2) and both biological and social risk factors play a relevant role in its onset and outcome. Psychopharmacology and psychotherapy are well-established and effective therapeutic options. (3) Thus, the risk of comorbidity associated with anxiety is high, increasing the complexity and challenge of its management. (1, 4, 5) Moreover, anxiety is a risk factor for suicidal thoughts and attempts, making it a serious public health challenge. (6)

Owing to social isolation, loss of loved ones, and medical and neuropsychiatric multimorbidity, notably cognitive decline, older adults are at a particularly high risk of experiencing anxiety. (7) Anxiety is ranked among the most prevalent mental health disorders in older adults, (2) and similar to other age groups, anxiety increases the risk of suicide in advanced ages. (8)

The comorbidity between thyroid dysfunction and affective disorders has long been studied and shown significant sex-dependent associations in different populations. (9) Depression is one of the main psychiatric disorders where thyroid hormones are significantly associated with its various diagnostic and therapeutic aspects. (10, 11) The underlying mechanisms are complex and anxiety was described as a common comorbidity in the association between thyroid dysfunction and depression. (12, 13) Furthermore, depression and thyroid dysfunction, mainly hypothyroidism, are also common disorders in advanced age groups. (2, 14, 15) Although the association between the hypothalamic-pituitary-thyroid axis and anxiety is well established, several studies did not adjust for depression as an eventual comorbidity. (16) Therefore, it is unclear whether thyroid function might be associated with anxiety independently from depression in older adults, and which factors might modulate the relationship.

The aim of this study was (1) to explore the association between thyroid function and anxiety in non-depressed older adults, (2) to assess the sex-related effect modification, and (3) to predict whether there is a non-linear pattern and relevant cutoff value in this association.

## 2. Methods

### 2.1. Study population

Alzheimer’s Disease Neuroimaging Initiative (ADNI) is a longitudinal naturalistic cohort study where aged adults with and without cognitive impairment or dementia were recruited across several centers in the United States and Canada. Study protocols, ethical approval, information, and data can be obtained from the official website https://adni.loni.usc.edu. Dr. Michael Weiner is the study’s principal investigator, which started in 2004 and is still ongoing. ADNI is funded as a private-public partnership with $27 million contributed by 20 companies and two foundations through the Foundation for the National Institutes of Health and $40 million from the National Institute on Aging. The study has five phases (ADNI 1, go, 2, 3, and currently 4). Different biomarkers of neurodegeneration are collected from included participants on a longitudinal course, mainly biological, genetic, neuroimaging, and neurocognitive. Some questionnaires were addressed to partners of study participants. All involved participants have given written consent. The study was performed according to the declaration of Helsinki and ethical approvals were obtained from the local IRBs corresponding to each recruitment site.

### 2.2. Thyroid function

The method was previously detailed in another study. (17) Thyroid function was estimated through serum Thyroid Stimulating Hormone (TSH) levels measured in µIU/dL (1 mIU/L=1 µIU/mL). TSH was assessed in fasting blood at baseline, and data was checked for completeness and correctness. Defect measurements were removed. Each set of duplicated measurements was individually compared to ensure consistency and chronology. When both are plausible, only the first value was retained ensuring the removal of inference related to eventual intervention between both dates. Very low, undetectable levels (< 0.01 µIU/mL) were reported in three cases and were converted to 0.01 µIU/mL allowing their inclusion in the statistical analyses. TSH of 10 µIU/mL and higher is the largely used cutoff value of subclinical hypothyroidism. (15) While the accuracy of the definition requires measurement of peripheral (free) thyroid hormone levels (Triiodothyronine: FT_3_ and Thyroxin: FT_4_), the two cases with TSH ≥ 10 µIU/mL were very high and therefore extracted from the dataset, and only cases with TSH < 10 µIU/mL were included in this study.

### 2.3. Anxiety symptoms

Anxiety symptoms were evaluated based on a relevant item of the neuropsychiatric inventory questionnaire (NPI or NPI-Q), where the study partner had to answer with “yes”, “no”, or “I don’t know”. Perceived anxiety was assessed as a binary variable (“yes= 1”, or “no= 0”), and “missing” or “I don’t know” answers were removed. The main anxiety-relevant NPI question is “Is the patient very nervous, worried, or frightened for no apparent reason? Does he/she seem very tense or fidgety? Is the patient afraid to be apart from you?”.

If this question was answered “yes”, depending on study-Phase, the following aspects might be further detailed.

1. “Does the patient say that he/she is worried about planned events?”
2. “Does the patient have periods of feeling shaky, unable to relax, or feeling excessively tense?”
3. “Does the patient have periods of (or complain of) shortness of breath, gasping, or sighing for no apparent reason other than nervousness?”
4. “Does the patient complain of butterflies in his/her stomach, or of racing or pounding of the heart in association with nervousness? (Symptoms not explained by ill health)”
5. “Does the patient avoid certain places or situations that make him/her more nervous such as riding in the car, meeting with friends, or being in crowds?”
6. “Does the patient become nervous and upset when separated from you (or his/her caregiver)? (Does he/she cling to you to keep from being separated?)”
7. “Does the patient show any other signs of anxiety?”

Only the binary answer to the main anxiety-related question was considered for the current analysis.

### 2.4. Neurocognitive tests and confounding factors

The Alzheimer’s Disease Assessment Score – 13 items (ADAS_13_) total score was included in the adjustment models to control for overall cognition. Moreover, the total scores of the Mini-Mental Status Examination (MMSE), Clinical Dementia Rating - Sum of Boxes (CDR-SB), and Functional Activities Questionnaire (FAQ) were also reported in the descriptive analysis as further neurocognitive biomarkers of the included cases. The total score of the Geriatric Depression Scale (GDS) was included to assess and exclude depression criteria.

Body-mass-index (BMI) was calculated as the result of “Weight (Kg) / (Height (m))*^2^*”. Cases missing one of those biometrical data or where the reported units did not match the given values and seemed erroneous were removed.

Sex (“Male” vs. “Female”), racial profile (“White”, “Black”, “Other”), cognition-related main diagnosis (“healthy control (HC)”, “Mild Cognitive Impairment (MCI)”, “dementia”), APOE ε4 (number of alleles: “none”, “one”, or “two”), educational level (in years), marital status (“currently married”, and “currently not married”: which includes “never married”, “widowed”, “divorced”, “unknown”), living condition/housing situation: “house or apartment”, “retirement or Nursing institution”, “other”), retirement (binary) were included as confounding factors.

### 2.5. Inclusion criteria

We included participants with valid TSH measurements, information on age or date of birth, defined baseline diagnosis, complete GDS scores and total GDS score ≤ 4 (exclusion of depression criteria), complete NPI or NPI-Q scores depending on the phase of ADNI in which the participant was initially recruited, complete ADAS_13_ score at baseline, and BMI.

After removing participants without TSH levels (n=159), without complete data on anxiety (n=22), without (complete) ADAS_13_ total scores at baseline (n=22), without (complete) GDS scores or where the total GDS score at baseline > 4 points (n=91), without information on age or baseline diagnosis (n=12), missing (n=4) or erroneous (n=6) biometric measurements, and those with TSH ≥ 10 µIU/mL (n=2), a total of 2,114 participants were included in this study. The study’s flow chart is detailed in Figure 1.

**Figure 1:**
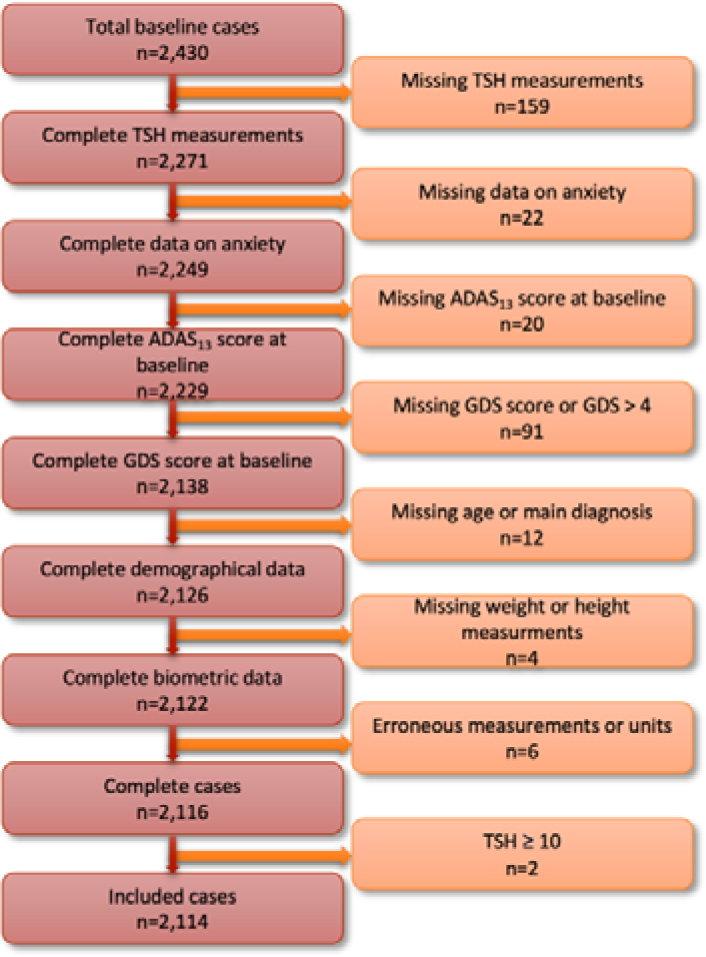
Chart flow of included cases.

### 2.6. Statistical analysis

Data analysis and visualization were performed by RStudio version 2024.04.1-748. After examination of the distribution, continuous variables were reported as median (IQR) and binary/categorical variables as number (%). Kruskal-Wallis rank sum test or Pearson’s Chi-squared test were used to compare groups of continuous or count data, respectively. The association between informant-reported anxiety and TSH was assessed with univariable logistic regression, where the dependent variable was the binary NPI/NPI-Q item on anxiety, and the independent variable was TSH level as a continuous variable (***<u>Model 1</u> – crude model***). Then relevant confounding factors were introduced in different models:

***Model 2 – demographic and biological factors:*** Model 1 + age + sex + racial profile + BMI.

**Model 3 – cognitive and social factors:** Model 2 + cognition-related main diagnosis + ADAS_13_ total score + educational level + APOE ε4 status + retirement status + housing situation + marital status.

The association was explored first in the total population, and then in each of the sex strata, after adapting the models (removing sex as a confounder).

A non-linear factor was introduced to the models and visualized using restricted cubic splines in the total study population and then sex strata. Based on a relevant knot value, a logistic regression analysis was performed comparing the two subgroups attributed to this cutoff value and adjusted for the same confounding factors corresponding to the previously mentioned models. Results were reported in OR, 95% CI, and p-value, and were considered statistically significant if the two-tailed p-value was ≤ 0.05.

## 3. Results

A total of 2,114 participants were included in this analysis, 773 (36.57%) were HC, 996 (47.11%) were classified as MCI, and 345 (16.32%) had dementia. The median age in the total population was 73 years (68–78), and 997 (47.16%) were females. The median ADAS_13_ baseline score was 14 points (9–22), the median GDS score was 1.00 point (0.00 – 2.00), and 290 (14%) had anxiety symptoms according to their study partner. The median TSH level was 1.69 µIU/mL (1.14 – 2.40). Details of the included population are summarized in Table 1. In the general population, TSH levels were significantly lower in those with anxiety than in those without (1.52 vs. 1.73 µIU/mL, p*_Mann-Whitney_*= 0.002). After stratification, the difference was statistically significant only in older males (1.40 vs. 1.76 µIU/mL, p*_Mann-Whitney_*<0.001)., but not older females. Comparative and correlation analyses of TSH values are visualized in Figure 2.

**Figure 2:**
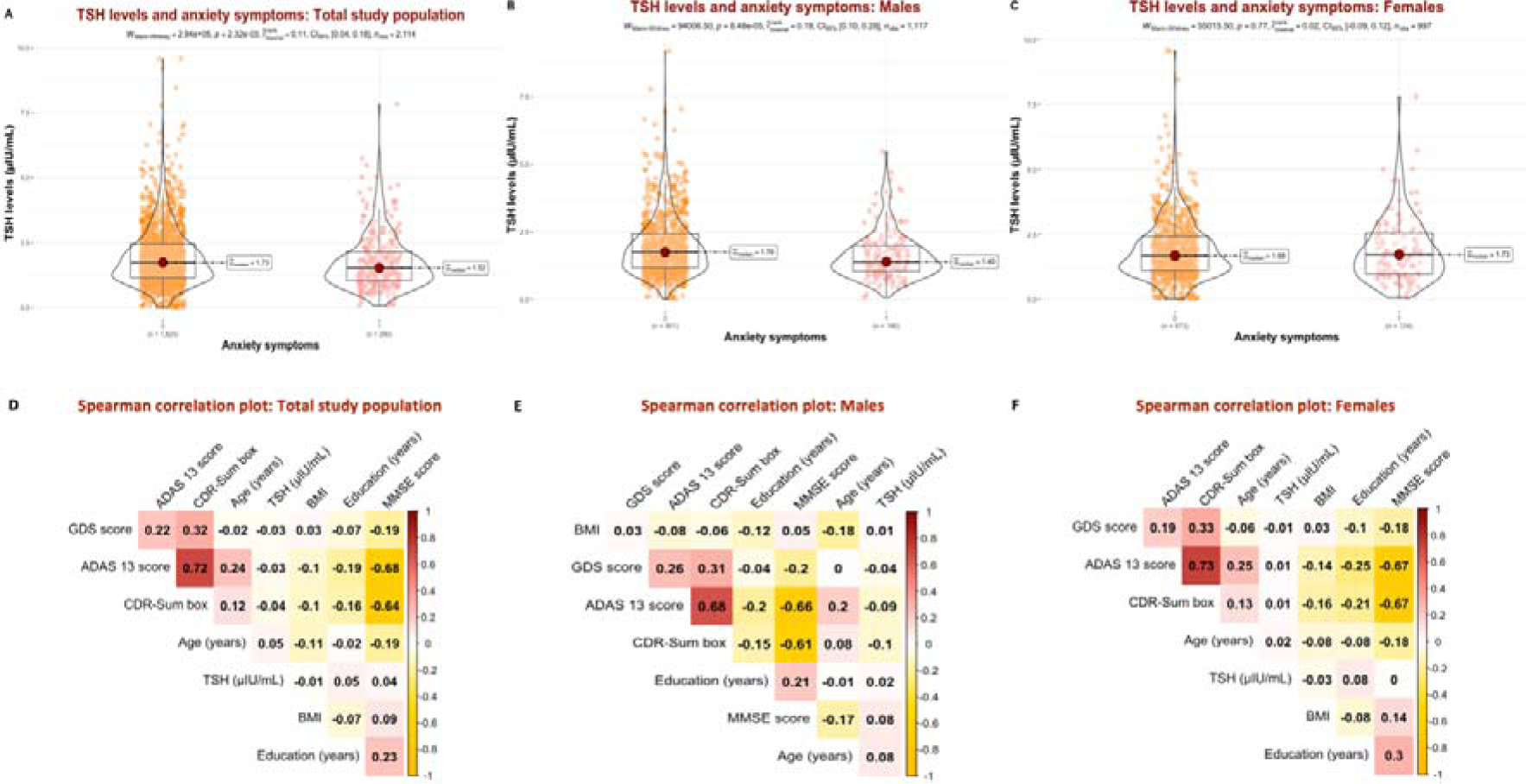
Thyroid-stimulating hormone (TSH) levels and associations. **2. A** C. omparative analyses of TSH depending on anxiety status in the total study population. **2. B.** Comparative analyses of TSH depending on anxiety status in the male stratum. **2. C.** Comparative analyses of TSH depending on anxiety status in the female stratum. **2. D.** Correlation analyses between thyroid stimulating hormone levels and biometric and neurocognitive data in the total study population. **2. E.** Correlation analyses between thyroid stimulating hormone levels and biometric and neurocognitive data in the male stratum. **2. F.** Correlation analyses between thyroid stimulating hormone levels and biometric and neurocognitive data in the female stratum.

**Table 1:**
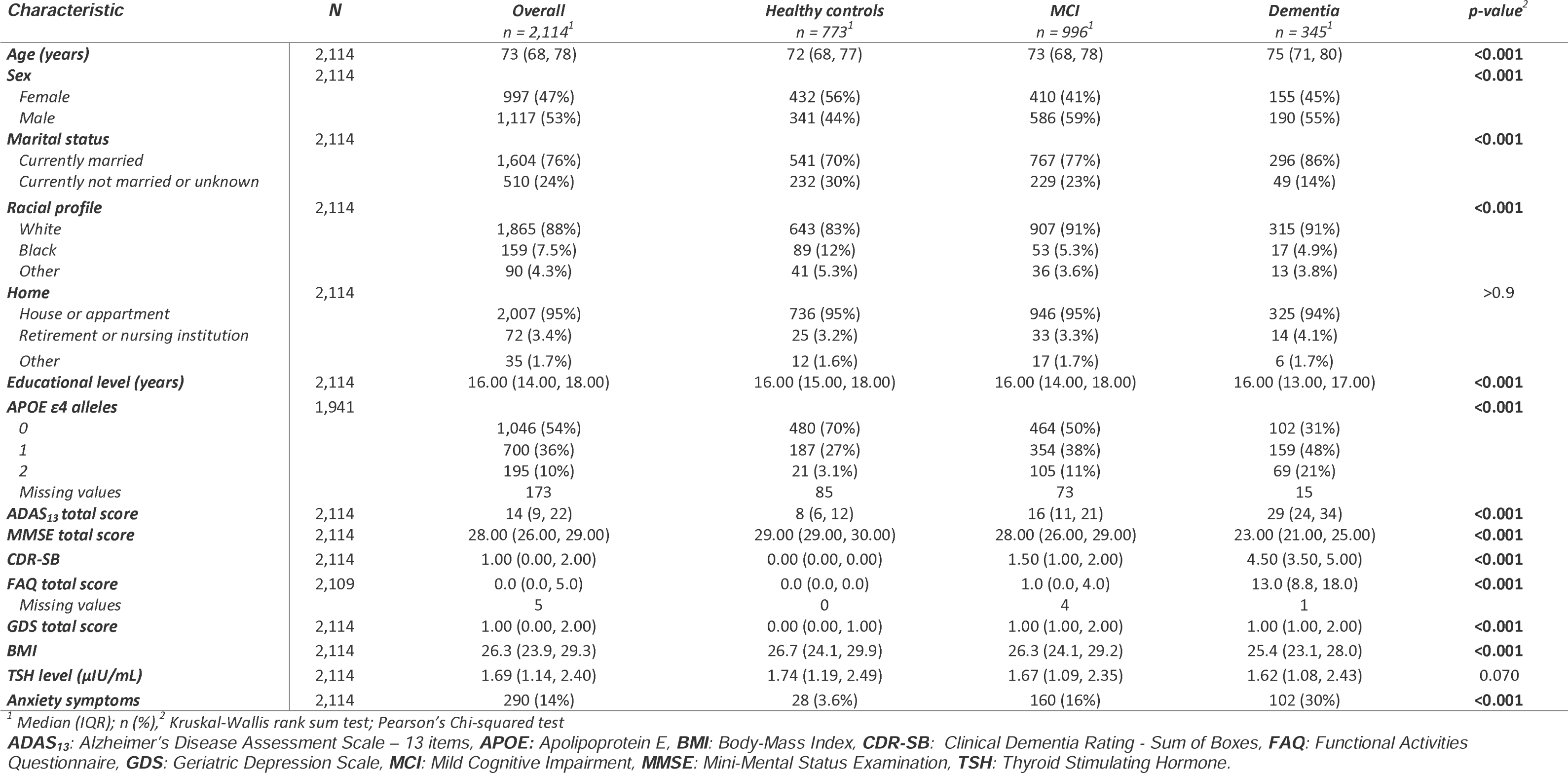
Characteristics of the study population.

| **Characteristic** | **N** | **Overall**<br>*n* = 2,114<sup>1</sup> | **Healthy controls**<br>*n* = 773<sup>1</sup> | **MCI**<br>*n* = 996<sup>1</sup> | **Dementia**<br>*n* = 345<sup>1</sup> | **p-value**<sup>2</sup> |
| --- | --- | --- | --- | --- | --- | --- |
| **Age (years)** | 2,114 | 73 (68, 78) | 72 (68, 77) | 73 (68, 78) | 75 (71, 80) | **<0.001** |
| **Sex** | 2,114 |  |  |  |  | **<0.001** |
| Female |  | 997 (47%) | 432 (56%) | 410 (41%) | 155 (45%) |  |
| Male |  | 1,117 (53%) | 341 (44%) | 586 (59%) | 190 (55%) |  |
| **Marital status** | 2,114 |  |  |  |  | **<0.001** |
| Currently married |  | 1,604 (76%) | 541 (70%) | 767 (77%) | 296 (86%) |  |
| Currently not married or unknown |  | 510 (24%) | 232 (30%) | 229 (23%) | 49 (14%) |  |
| **Racial profile** | 2,114 |  |  |  |  | **<0.001** |
| White |  | 1,865 (88%) | 643 (83%) | 907 (91%) | 315 (91%) |  |
| Black |  | 159 (7.5%) | 89 (12%) | 53 (5.3%) | 17 (4.9%) |  |
| Other |  | 90 (4.3%) | 41 (5.3%) | 36 (3.6%) | 13 (3.8%) |  |
| **Home** | 2,114 |  |  |  |  | >0.9 |
| House or apartment |  | 2,007 (95%) | 736 (95%) | 946 (95%) | 325 (94%) |  |
| Retirement or nursing institution |  | 72 (3.4%) | 25 (3.2%) | 33 (3.3%) | 14 (4.1%) |  |
| Other |  | 35 (1.7%) | 12 (1.6%) | 17 (1.7%) | 6 (1.7%) |  |
| **Educational level (years)** | 2,114 | 16.00 (14.00, 18.00) | 16.00 (15.00, 18.00) | 16.00 (14.00, 18.00) | 16.00 (13.00, 17.00) | **<0.001** |
| **APOE ε4 alleles** | 1,941 |  |  |  |  | **<0.001** |
| 0 |  | 1,046 (54%) | 480 (70%) | 464 (50%) | 102 (31%) |  |
| 1 |  | 700 (36%) | 187 (27%) | 354 (38%) | 159 (48%) |  |
| 2 |  | 195 (10%) | 21 (3.1%) | 105 (11%) | 69 (21%) |  |
| Missing values |  | 173 | 85 | 73 | 15 |  |
| **ADAS<sub>13</sub> total score** | 2,114 | 14 (9, 22) | 8 (6, 12) | 16 (11, 21) | 29 (24, 34) | **<0.001** |
| **MMSE total score** | 2,114 | 28.00 (26.00, 29.00) | 29.00 (29.00, 30.00) | 28.00 (26.00, 29.00) | 23.00 (21.00, 25.00) | **<0.001** |
| **CDR-SB** | 2,114 | 1.00 (0.00, 2.00) | 0.00 (0.00, 0.00) | 1.50 (1.00, 2.00) | 4.50 (3.50, 5.00) | **<0.001** |
| **FAQ total score** | 2,109 | 0.0 (0.0, 5.0) | 0.0 (0.0, 0.0) | 1.0 (0.0, 4.0) | 13.0 (8.8, 18.0) | **<0.001** |
| Missing values |  | 5 | 0 | 4 | 1 |  |
| **GDS total score** | 2,114 | 1.00 (0.00, 2.00) | 0.00 (0.00, 1.00) | 1.00 (1.00, 2.00) | 1.00 (1.00, 2.00) | **<0.001** |
| **BMI** | 2,114 | 26.3 (23.9, 29.3) | 26.7 (24.1, 29.9) | 26.3 (24.1, 29.2) | 25.4 (23.1, 28.0) | **<0.001** |
| **TSH level (μIU/mL)** | 2,114 | 1.69 (1.14, 2.40) | 1.74 (1.19, 2.49) | 1.67 (1.09, 2.35) | 1.62 (1.08, 2.43) | 0.070 |
| **Anxiety symptoms** | 2,114 | 290 (14%) | 28 (3.6%) | 160 (16%) | 102 (30%) | **<0.001** |

<sup>1</sup> Median (IQR); *n* (%), <sup>2</sup> Kruskal-Wallis rank sum test; Pearson's Chi-squared test
**ADAS<sub>13</sub>**: Alzheimer's Disease Assessment Scale – 13 items, **APOE**: Apolipoprotein E, **BMI**: Body-Mass Index, **CDR-SB**: Clinical Dementia Rating - Sum of Boxes, **FAQ**: Functional Activities Questionnaire, **GDS**: Geriatric Depression Scale, **MCI**: Mild Cognitive Impairment, **MMSE**: Mini-Mental Status Examination, **TSH**: Thyroid Stimulating Hormone.

The regression analysis showed a significant association between anxiety and TSH levels in the main study population (OR_Model1_= 0.86, 95% CI: 0.76 – 0.97, p-value_Model1_= 0.011), even after adjusting for bio-demographical (adj. OR_Model2_= 0.85, 95% CI: 0.75 – 0.96, p-value_Model2_=0.007), and socio-cognitive confounders (adj. OR_Model3_= 0.84, 95% CI: 0.73 – 0.96, p-value_Model3_= 0.009). The sex stratification showed a significant association between anxiety and TSH levels in male participants (OR_Model1-male_= 0.71, 95% CI: 0.58 – 0.85, p-value_Model1-male_<0.001), but not in females (OR _Model1-female_= 1.01, 95% CI: 0.86 – 1.18, p-value_Model1-female_= 0.878). After adjusting for different variables, the results remained consistent in the male groups (adj. OR_Model2-male_= 0.71, 95% CI: 0.59 – 0.86, p-value_Model2-male_<0.001 and adj. OR_Model3-male_= 0.71, 95% CI: 0.57 – 0.86, p-value_Model3-male_<0.001). No statistical significance was found in the female stratum using different models. Logistic regression results based on the continuous TSH as a predictor variable are summarized in Supplementary Table 1.

Introducing cubic restricted splines to the models showed a non-linear pattern. The 3 quartile (75 percentile) corresponding to a TSH value of 2.4 µIU/mL was a significant cutoff value, under which the association between TSH and OR of anxiety was particularly statistically significant in the main population and male stratum (Figure 3). Moreover, participants with TSH ≤ 2.4 µIU/mL had 35% higher odds of perceived anxiety than those with TSH > 2.5 µIU/mL in the main study population (OR_Model1_= 1.35, 95% CI: 1.00 – 1.85, p-value*_Model1_*= 0.048). The association remained significant after adjusting for bio-demographical (OR_Model2_= 1.37, 95% CI: 1.02 – 1.88, p-value*_Model2_*= 0.039) and socio-cognitive confounding factors (OR_Model3_= 1.48, 95% CI: 1.06 – 2.10, p-value*_Model3_*= 0.021).

**Figure 3:**
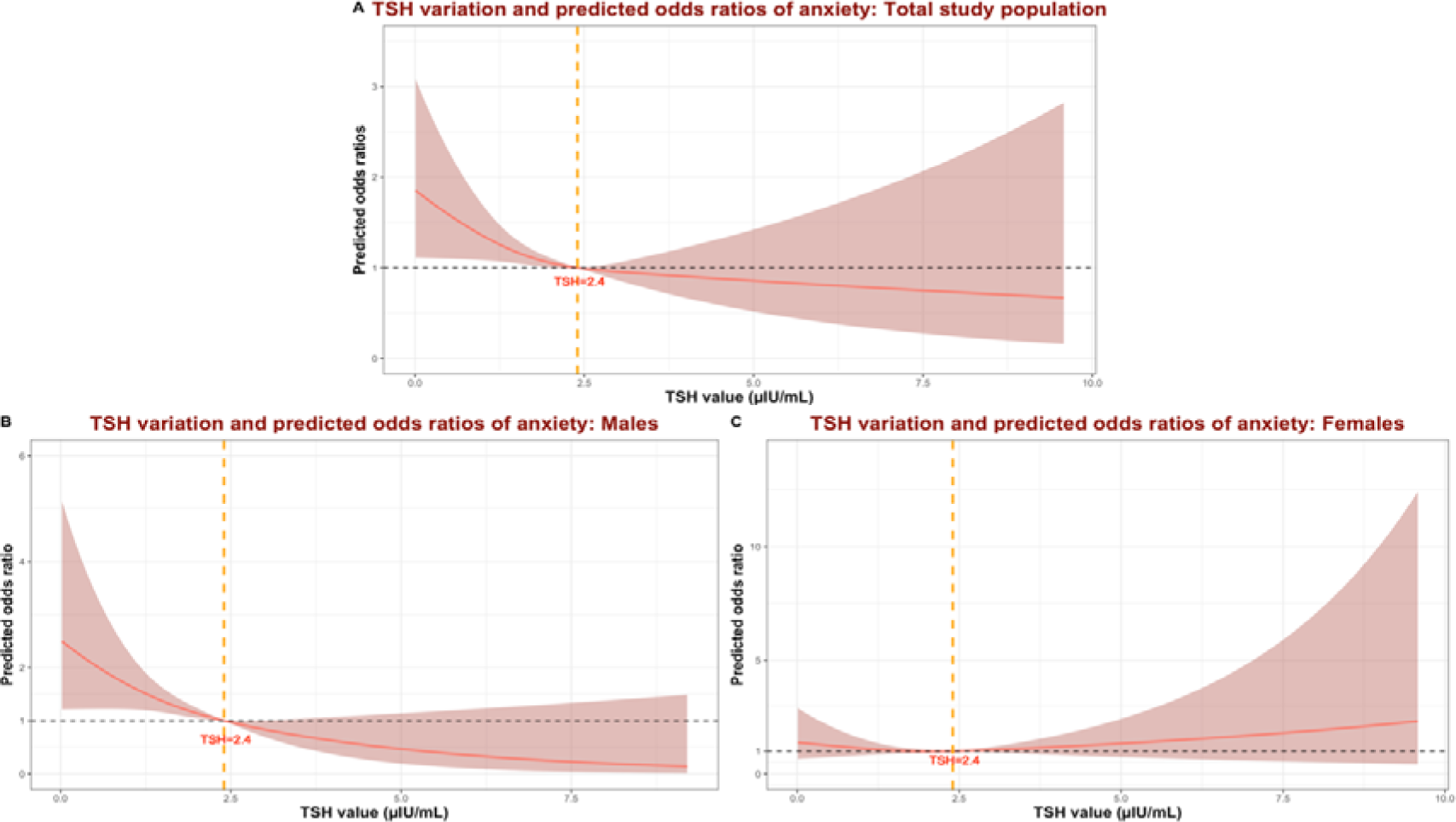
Predicted odds ratios of anxiety depending on thyroid-stimulating hormone levels in the total study population and sex strata.

The association was stronger in the male stratum, where participants with TSH ≤ 2.4 µIU/mL had 128% higher odds of perceived anxiety than those with TSH > 2.4 µIU/mL (OR_Model1-male_= 2.28, 95% CI: 1.45 – 3.75, p-value*_Model1-male_*<0.001). Similar findings were found after adjusting for relevant confounding factors in models 2 and 3. No Significant associations were found in the female stratum. Logistic regression results based on the categorical TSH as a predicting variable are summarized in Supplementary Table 2.

A summary overview of different prediction models is visualized in Figure 4.

**Figure 4:**
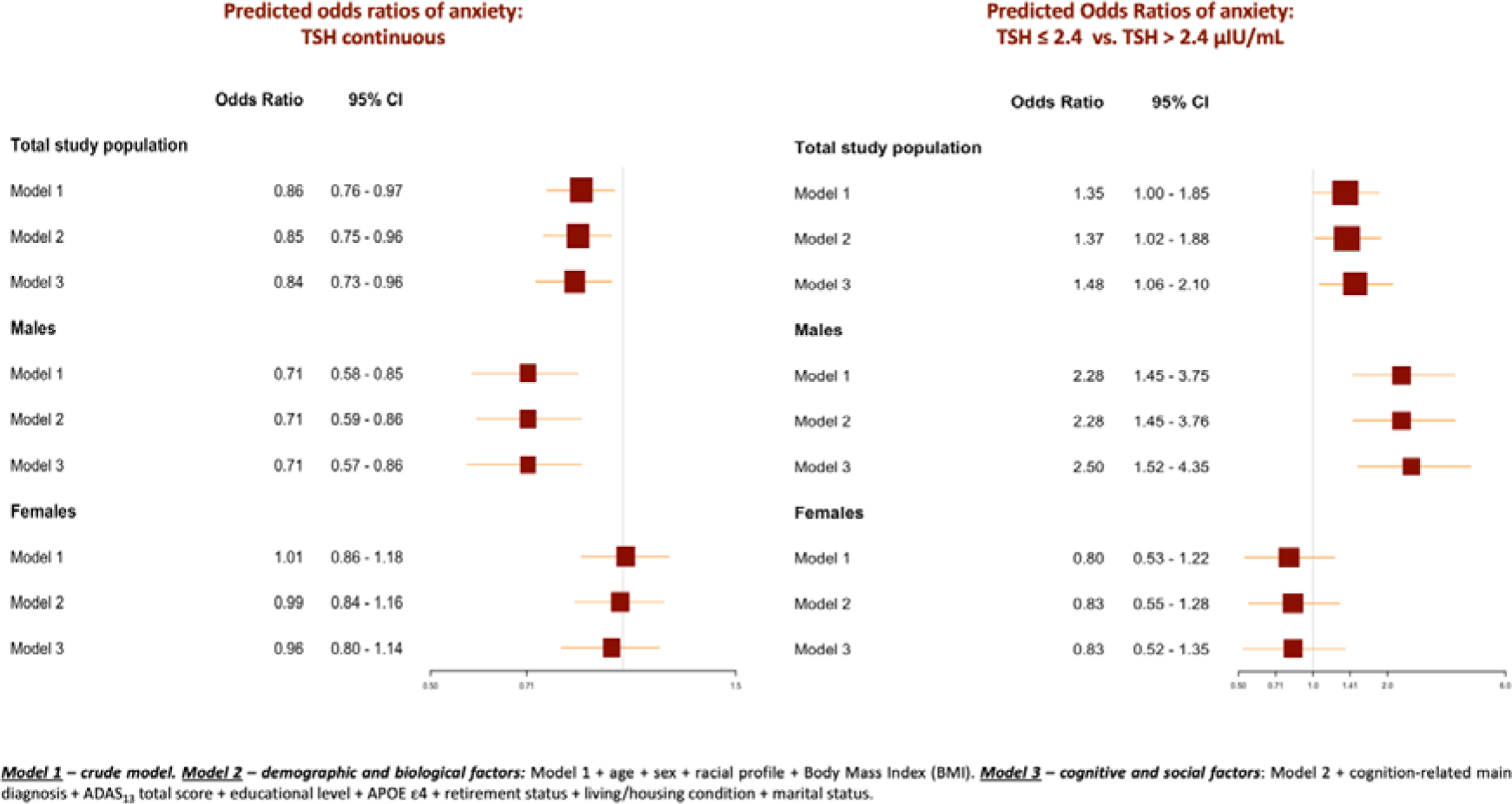
Summary Forest plot of odds ratios of anxiety depending on thyroid function in the total study population and sex strata and comparison of different prediction models.

## 4. Discussion

This study explored the association between thyroid function and anxiety symptoms in older adults, as perceived and reported by their study partners.

The main outcome was the significant association between TSH levels and perceived anxiety in non-depressed males, even after adjustment for different confounding factors such as age, race, BMI, presence of MCI or dementia, educational level, overall cognition, and social parameters (marital status, housing situation, and retirement status). Higher TSH levels were significantly associated with lower odds of reporting anxiety symptoms in male participants. A TSH level of 2.4 µIU/mL was a relevant cutoff value for this association. This significant association was also found in the main population but not in the stratum of non-depressed older female participants. Sex-modulated TSH effect on anxiety symptoms beyond depression was rarely studied, and most of the reported studies explored associations in patients with major depression as the main diagnosis.

### 4.1. Associations between TSH and anxiety

In young drag naïve patients with the first-episode major depressive episode, those with subclinical hypothyroidism had a higher prevalence of anxiety (15.8%). Higher TSH levels predicted a higher prevalence of anxiety and a TSH level of 6.17 mIU/L (equal to 6.17 µIU/mL) was a significant cutoff point in distinguishing patients with anxiety symptoms from those without. (18) Similar results were reported in further studies, where higher TSH levels predicted anxiety in patients with first-episode major depressive episodes. (12, 13)

Higher TSH levels were significant predictors of suicide attempts in patients with first-episode major depressive disorder (19) particularly in young males. (20) In severely depressive patients, thyroid function was also associated with psychotic symptoms. (21) Both anxiety and high TSH levels were significant mediators in the association between depression and psychotic symptoms. (22)

The significant associations with high TSH levels reported in these studies contradict our findings, and several major differences between the different study populations might explain the discrepancies: first, the included participants in the current study had no depression symptoms, in contrast with the findings of reported studies, where hypothyroidism might modulate the severity of depression, inducing more fatigue, less motivation, cognitive slowing, and consequently higher risks of anxiety symptoms as comorbidity. (12, 13) Second, TSH levels tend to increase physiologically with age in older adults, and the sensitivity of the nervous system might vary across age groups. Here only older adults were studied, in contrast with published data.

Several studies reported significant interactions between metabolic syndrome (or blood glucose levels), thyroid hormones, and anxiety. (23–28) Therefore, our prediction models were also adjusted for BMI. Although a significant effect of BMI was seen in Model 2, the statistical significance was lost following the introduction of socio-cognitive confounders in Model 3. Overall, the association between TSH and anxiety remained unaffected by the BMI.

### 4.2. Sex-related differences

The association of thyroid hormones with affective disorders was mediated by sex in different populations and age groups. In one study on patients with major depressive episodes, TSH and thyroid-specific antibodies predicted anxiety only in males, (29) while the association between anti-thyroglobulin antibodies and anxiety was significant in females in another study. (30) However, autoimmune-mediated thyroid pathologies are more common in women, (31) and this might explain some inconsistencies in the different results depending on the studied population.

In a study where only men of different age groups were included, thyroid disorders were more prevalent in those with anxiety than those without (6.7% vs. 1.9%, p-value= 0.016), and predicted higher odds of anxiety disorders (adj. OR= 5.54, 95% CI: 1.64 - 18.64, p-value= 0.006). The association was not significant when testing for mood disorders. (32)

The observed absence of an association between TSH and anxiety in older females does not exclude significant associations in younger age groups and different circumstances. A longitudinal study in euthyroid pregnant women showed that thyroid hormone variations were associated with depression, anxiety, and obsessive-compulsive disorder scores. (33)

### 4.3. Hypotheses

While there were several studies on the different effects of thyroid hormones on neuropsychiatric and affective disorders, various underlying mechanisms might be involved. Thyroid hormones modulate brain metabolism, particularly in emotion- and cognition-regulating brain structures. Hypothyroidism was associated with reduced regional glucose uptake, and the anomalies were reversible after thyroid hormone substitution. (34) Thyroid hormones modulate the release and regulation of brain neurotransmitters, (35, 36) inflammatory biomarkers, (37, 38) and cerebral gene expression. (39, 40) Furthermore, N-methyl-D-Aspartate (NMDA) receptors were found to be involved in the association between anxiety and thyroid lesions. (41)

The difference in the TSH level-anxiety association between males and females is less understood. One hypothesis might be supported by the theory that males and females exhibit distinct emotional expression patterns. (42) The results of the current study were based on an external perception of anxiety in study participants and might therefore be biased by this aspect. Another hypothesis supports the role of gonadal hormones in mediating the effect of thyroid hormones on the brain. (43, 44)

### 4.4. Limitations

The main limitation of our study is the absence of data on peripheral thyroid hormones such as FT_3_ and FT_4_, both of which have an inverse relationship with TSH levels. Although these measurements give additional information on the peripheral thyroid function, they were not accessed in the accessible ADNI baseline laboratory investigation. The focus of this study was TSH as a biomarker of thyroid function and as an exposure in the statistical models. Extreme values were either replaced for the statistical analysis (three cases with TSH < 0.01 µIU/mL) or removed (two cases with TSH ≥ 10 µIU/mL).

The second limitation is the lack of information on prior thyroid affections such as Grave’s disease, Hashimoto thyroiditis, or postpartum thyroiditis. These are associated with further autoimmunity risk factors and with additional comorbidities, leading to higher anxiety levels and depression. (45–47) Etiologies were kept out of the scope of the current studies owing to the variability of autoimmune disorders that might affect TSH levels, (48) and to avoid recall and information bias associated with reporting non-cognition-specific comorbidities in ADNI. TSH was a preferred biomarker for this study. In a large study on 1,017 youth admitted for psychiatric disorders, 6% presented abnormal TSH levels, while only less than 1% had thyroid pathologies. (49) The lack of information on whether participants might have had, or currently have, thyroid cancers or adenomas might be a further limitation since oncologic comorbidity might increase the predisposition for anxiety. (50, 51) The exclusion of participants with depression was a way to adjust for this eventual confounding factor. Moreover, ADNI inclusion criteria were strict, and a severe concomitant active neoplasm might have disqualified the participant.

A further limitation is the absence of information on whether study participants have had thyroid ablation therapy and are under thyroid hormone supplementation. While this information might help draw an overall prevalence of underlying thyroid pathologies and understand the etiology of pathological TSH levels, it is unexpected that this missing information might affect current results regarding anxiety symptoms in the context of TSH variation.

Finally, anxiety was assessed based on a questionnaire answered by study partners and anxiety symptoms were addressed in only one question. Therefore, replicating the study using conventional anxiety screening tools, such as the Hamilton Anxiety Rating Scale (HAM-A), is important.

## 5. Conclusions

The study suggests a sex-modulated association between thyroid function and informant-perceived anxiety in non-depressed older populations. This presents another argument on an interrelation between the brain and the thyroid, independently from the commonly explored depression. A TSH level of 2.4 µIU/mL was a relevant cutoff value for the association, and higher odds of reported anxiety were observed in the lower range compared to the higher range. Informant-reported anxiety might be a biomarker of low TSH levels, and vice versa, and need to motivate further investigations. The differences in results between older males and females are thus still not well understood. More studies are needed in order to assess the sex-related biological predisposing and confounding factors.

## Declarations

### Data availability

All data used in the manuscript is available at https://adni.loni.usc.edu. **Consent for publication:** Brain and Thyroid vectors used in the graphical abstract are designed by Freepik (License free).

### Author contributions

AH has full access to all of the data and takes responsibility for the integrity of the data and the accuracy of the analysis, visualization, drafting, and editing of the manuscript.

*Data used in preparation of this article were obtained from the Alzheimer’s Disease Neuroimaging Initiative (ADNI) database (adni.loni.usc.edu). As such, the investigators within the ADNI contributed to the design and implementation of ADNI and/or provided data but did not participate in analysis or writing of this report. A complete listing of ADNI investigators can be found at: http://adni.loni.usc.edu/wp-content/uploads/how_to_apply/ADNI_Acknowledgement_List.pdf

## Acknowledgments

“Data collection and sharing for the Alzheimer’s Disease Neuroimaging Initiative (ADNI) is funded by the National Institute on Aging (National Institutes of Health Grant U19 AG024904). The grantee organization is the Northern California Institute for Research and Education. In the past, ADNI has also received funding from the National Institute of Biomedical Imaging and Bioengineering, the Canadian Institutes of Health Research, and private sector contributions through the Foundation for the National Institutes of Health (FNIH) including generous contributions from the following: AbbVie, Alzheimer’s Association; Alzheimer’s Drug Discovery Foundation; Araclon Biotech; BioClinica, Inc.; Biogen; Bristol-Myers Squibb Company; CereSpir, Inc.; Cogstate; Eisai Inc.; Elan Pharmaceuticals, Inc.; Eli Lilly and Company; EuroImmun; F. Hoffmann-La Roche Ltd and its affiliated company Genentech, Inc.; Fujirebio; GE Healthcare; IXICO Ltd.; Janssen Alzheimer Immunotherapy Research & Development, LLC.; Johnson & Johnson Pharmaceutical Research &Development LLC.; Lumosity; Lundbeck; Merck & Co., Inc.; Meso Scale Diagnostics, LLC.; NeuroRx Research; Neurotrack Technologies; Novartis Pharmaceuticals Corporation; Pfizer Inc.; Piramal Imaging; Servier; Takeda Pharmaceutical Company; and Transition Therapeutics.”

---

**Supplementary Table 1:**
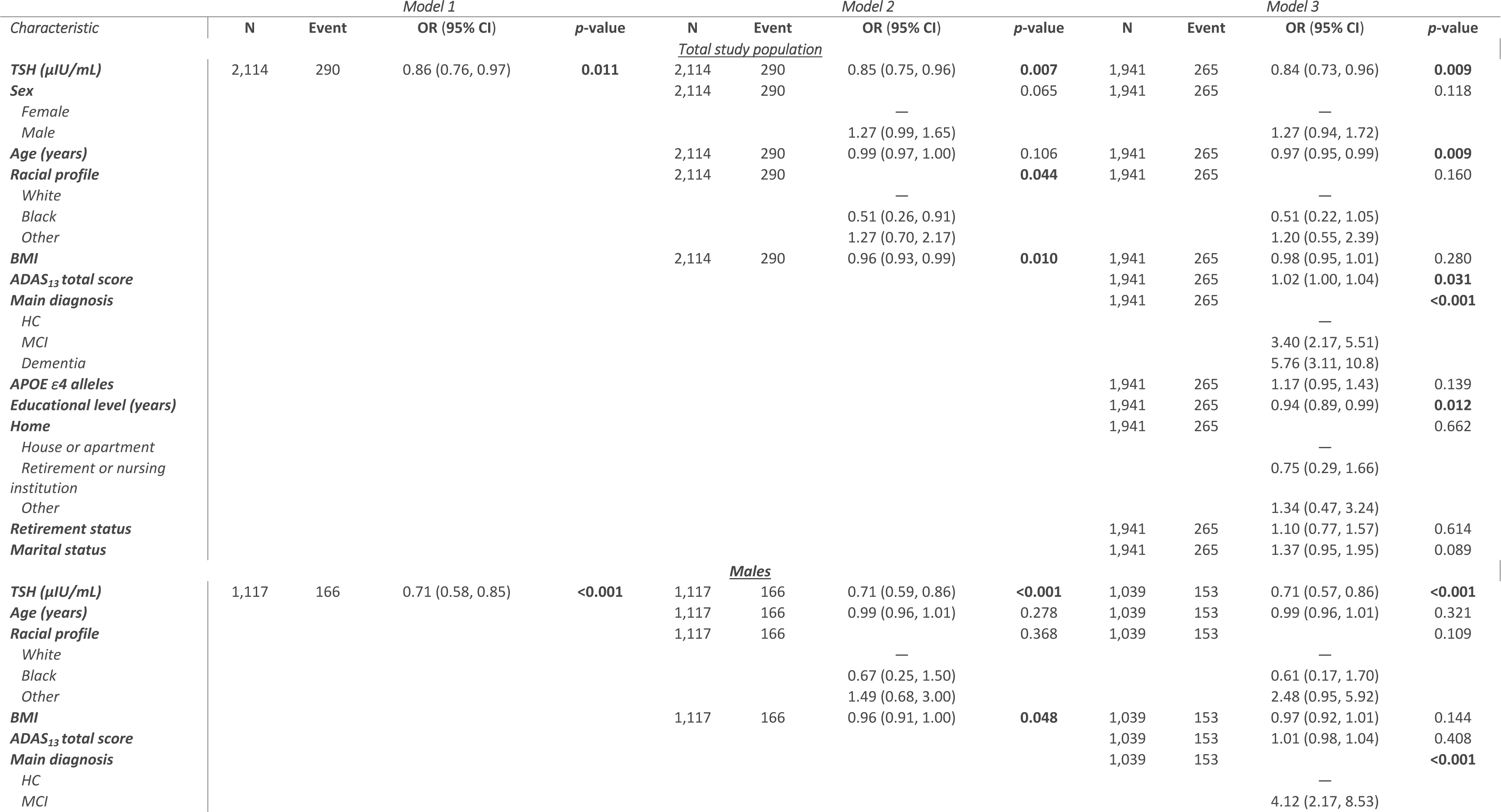
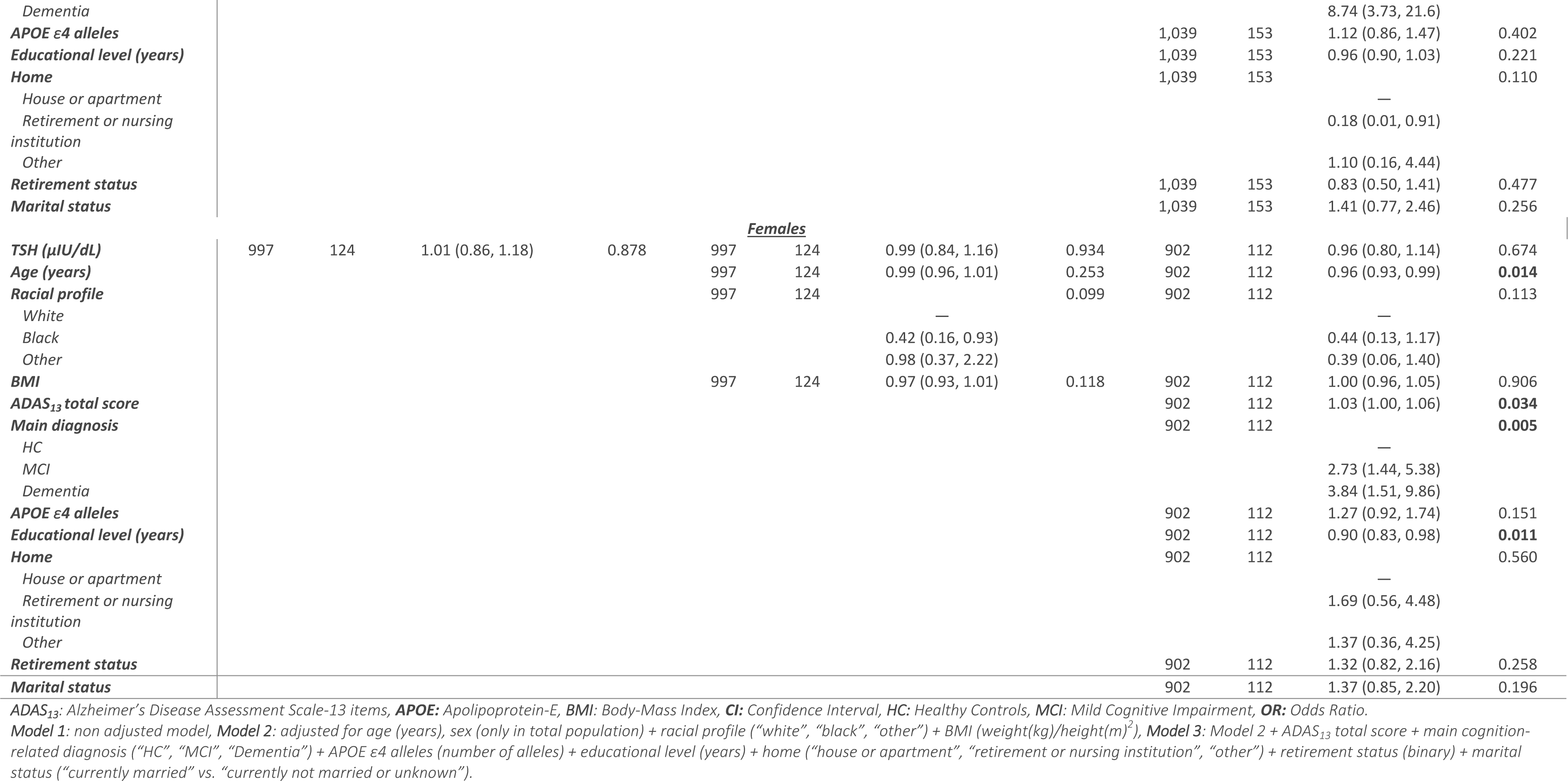
Detailed models of the association between thyroid function (TSH continuous) and anxiety symptoms in the total population and sex strata.

**Supplementary Table 2:**
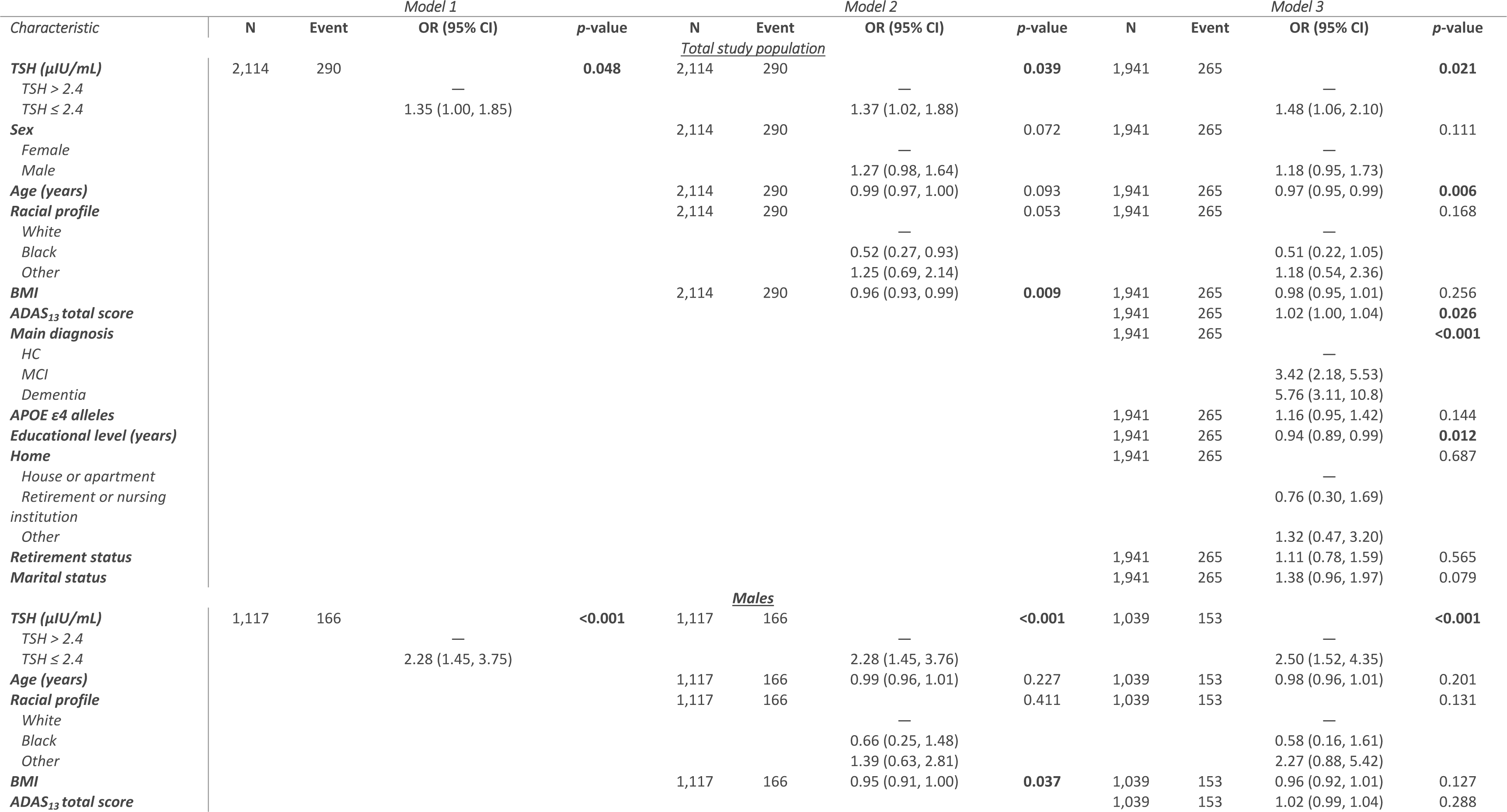
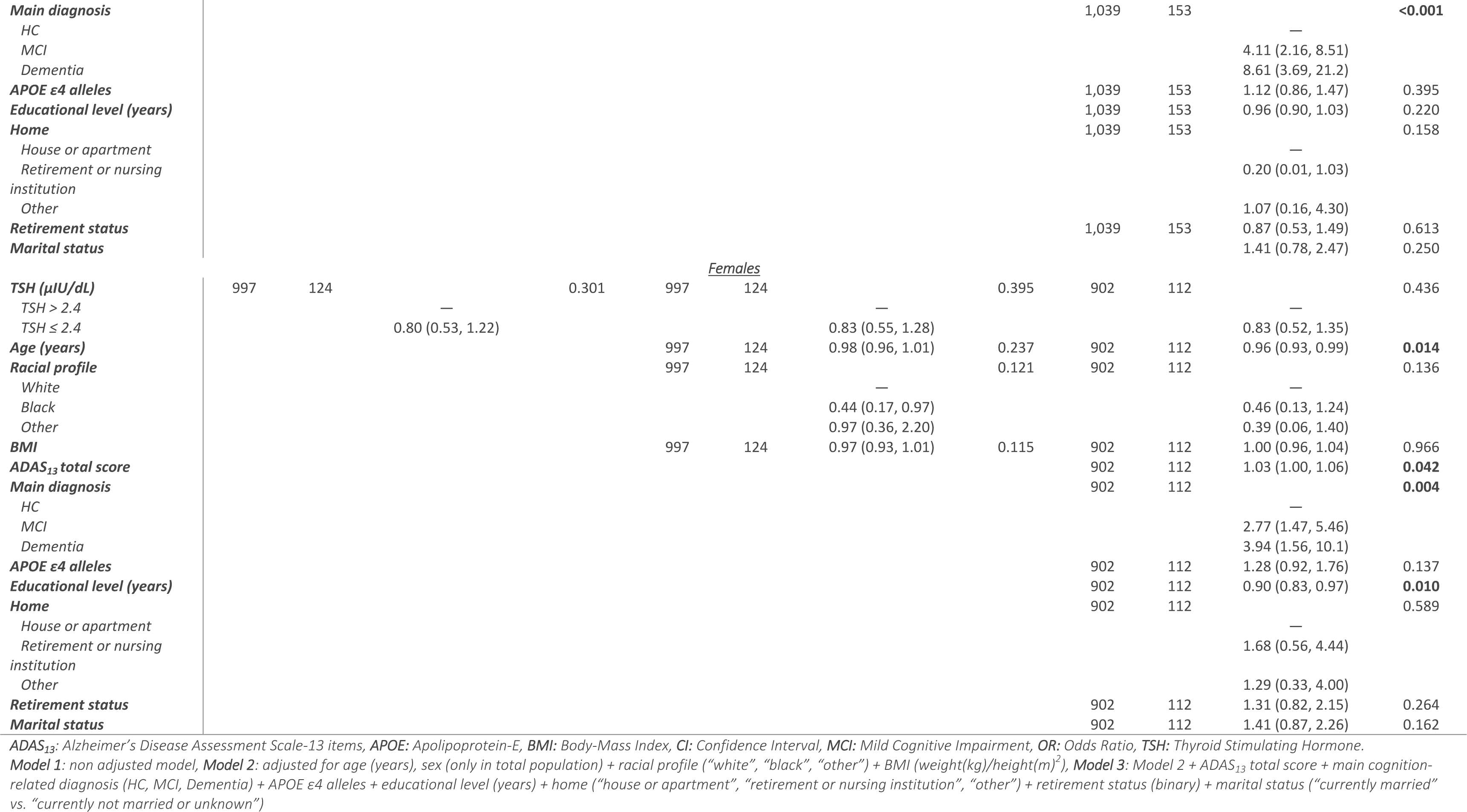
Detailed models of the association between thyroid function (TSH categorical) and anxiety symptoms in the total population and sex strata.

